# Attractive Targeted Sugar Baits for malaria control in western Kenya (ATSB-Kenya) – Effect of ATSBs on epidemiologic and entomologic indicators: a Phase III, open-label, cluster-randomised, controlled trial

**DOI:** 10.1101/2025.01.14.25320559

**Authors:** Caroline Ogwang, Aaron M Samuels, Daniel P McDermott, Alice Kamau, Maia Lesosky, Kizito Obiet, Julia M Janssen, Wycliffe Odongo, John E Gimnig, Julie R Gutman, Jonathan S Schultz, Oliver Towett, Brian Seda, Mercy Chepkirui, Margaret Muchoki, Seline Omondi, Jackline Kosgei, Brian Polo, Frank Aduwo, Kephas Otieno, Martin J Donnelly, Simon Kariuki, Eric Ochomo, Feiko ter Kuile, Sarah G Staedke

**Author notes:** Corresponding author; (CO). Current Address: National Heart and Lung Institute, Imperial College London, UK. These authors contributed equally to this work (Feiko ter Kuile, Sarah G Staedke).

## Abstract

Attractive targeted sugar baits (ATSBs) are a novel malaria control tool designed to target mosquitoes outdoors. We conducted a cluster-randomised trial to evaluate the impact of ATSBs on malaria indicators in Kenya. Seventy clusters (≥100 households/cluster) in Siaya county were randomly assigned (1:1) to intervention or control. Pyrethroid-only long-lasting insecticidal nets were distributed to all clusters, aiming for universal coverage. Two ATSBs containing dinotefuran were hung outside household structures in intervention clusters. ATSBs were monitored every two months and replaced every six months over two years. Three consecutive cohorts of randomly selected children (1-<15 years) were enrolled, aiming to accrue 1,260 person-years over two years of follow-up. Incidence of clinical malaria (fever with a positive malaria test) was the primary outcome. A multilevel Poisson regression model was applied, with clusters as a random intercept and study arm as a fixed effect. Secondary outcomes were malaria prevalence in community residents (≥1 month), and parity of mosquitos captured through human landing catches. In March 2022, ATSBs were delivered to 33,180 of 33,419 (99.3%) household structures in intervention clusters. Overall, 268,268 ATSBs were deployed over two years. Of 2,962 cohort children enrolled (intervention=1,497; control=1,465), 2,869 (96.9%) were included in the primary analysis (intervention=1,461; control=1,408), contributing 1,445 person-years of follow-up. Malaria incidence was 1.32 episodes per person-years in the intervention arm versus 1.20 in the control (unadjusted incidence rate ratio 1.11; 95% CI: 0.75-1.65; p=0.598). Of 7,488 community residents surveyed(intervention=3,760; control=3,728), 1,474 (39.2%) intervention and 1,461 (39.2%) control participants tested positive for malaria (unadjusted odds ratio [OR] 0.98; 95% CI: 0.60-1.59; p=0.93). Of 6,457 female anopheles mosquitoes collected (intervention=4,058; control=2,399), 3,579 (88.2%) intervention and 1,973 (82.2%) control mosquitoes were parous (OR 1.34; 95% CI: 0.91-1.99; p=0.14). In Kenya, we found no evidence that ATSBs reduced clinical malaria incidence, malaria prevalence, or vector parity.

**Trial registration:** Clinicaltrials.gov (NCT05219565), 22 January 2022; https://clinicaltrials.gov/study/NCT05219565

## Introduction

Between 2000 to 2015, the prevalence and incidence of *Plasmodium falciparum* in Africa fell by 50% and 40%, respectively, primarily due to the scale-up of long-lasting insecticidal nets (LLINs), indoor residual spraying (IRS) and treatment with artemisinin-based combination therapies (ACTs) [1]. Since 2015, however, progress on malaria control has plateaued [2]. Malaria control efforts were hindered by the COVID-19 pandemic, but widespread resistance to pyrethroid insecticides, climate change, fluctuations in vector species, changes in mosquito behaviours, and resistance to artemisinin and partner drugs are other threats [2–6]. In Kenya, malaria burden remains high in the western, lake- endemic region, despite distribution of LLINs and targeted IRS [7].

LLINs are the most widely used vector control tool, supplemented by IRS [2]. Both tools target mosquito vectors that feed and rest indoors. However, vector behaviours are shifting, with increased biting outdoors and later in the morning hours, when individuals are less likely to be protected by LLINs [5, 8]. Novel tools, such as attractive targeted sugar baits (ATSBs) that target mosquitoes outdoors are needed to complement other control interventions deployed indoors. ATSBs are A4- sized panels containing fruit syrup laced with an insecticide, which exploits the sugar-feeding behaviour of mosquitoes [9]. ATSBs attract mosquitoes, luring them to feed and ingest the insecticide, which kills the mosquitoes.

In 2017, an initial entomological field trial of ATSBs was conducted in Mali, which found ATSBs were associated with significant reductions in vector density, sporozoite rate, and entomologic inoculation rate (EIR) associated with ATSBs [10]. Subsequent modelling studies suggested that ATSBs could reduce malaria incidence and parasite prevalence by over 30% annually [11]. Given these promising results, results from Zambia indicating that *Anopheles funestus* and *An. gambiae* fed readily from the ATSB stations [12], and the predicted impact of ATSBs on malaria transmission [11, 13], large-scale, Phase III cluster-randomised, controlled trials were launched in 2021 to further evaluate the epidemiological and entomological impact of ATSBs in Kenya, Mali and Zambia [14]. Here, we present the results of the Kenyan trial.

## Methods

The protocol and statistical analysis plan for the multi-country ATSB trial have been published previously [14, 15]. A description of cohort recruitment (Fig1), and the characteristics of children and households at enrolment, have also been published [16].

### Study setting

The trial was conducted in Alego-Usonga and Rarieda sub-counties in Siaya County, western Kenya (Fig2), in the region of the Health and Demographic Surveillance System (HDSS) established by the Kenya Medical Research Institute and the US Centers of Disease Control and Prevention, which has been described in detail [17, 18]. Briefly, malaria transmission is perennial in this area with prevalence peaking between May-June and November-December, following the long and short rains. Malaria prevalence by microscopy in the study area was 36.8% in 2013, and was highest in children aged 5-14 years (57.9%) [19]. The primary malaria vectors are *An. funestus* and *An. arabiensis* [5, 20]. Artemether-lumefantrine (AL) has been used as the first-line therapy for case management since 2006, LLIN mass campaigns are conducted every 3-5 years (2014, 2017, 2021), and IRS is not implemented in this area [21].

### Assignment of interventions

Following a census conducted from December 2020 to March 2021, we established 80 clusters by purposely grouping 1-3 adjacent villages to create a cluster of sufficient population size. To minimise contamination, clusters were divided into core and buffer areas, with core areas 300-600 meters from the perimeter, and 600-1,200 meters between cores of contiguous clusters. We aimed for a core population of 100-400 households. Prior to randomisation, malaria incidence, prevalence, and contextual data were collected in the 80 clusters. Subsequently, 10 clusters with the fewest children aged 1-<15 years were removed, to limit resampling of individuals. Thus, 70 of the original 80 clusters were included in the trial.

Clusters were assigned to study arms using restricted randomisation to minimise imbalances, considering key baseline characteristics (malaria incidence, bednet use, RTS,S/AS01 vaccination implemented within the cluster, housing density, study clinic location, altitude, and entomological indicators), as described previously [14, 15]. A list of 100,000 possible allocation sequences was generated by a study team member not involved in trial implementation and checked against balance criteria. Any sequences that failed to balance these criteria were dropped. The remaining sequences were assessed for validity, and one was chosen randomly. Assignment to intervention or control was determined by a coin toss. The 70 clusters (32 in Alego Usonga and 38 in Rarieda) were randomised in a 1:1 ratio, 35 to the intervention arm and 35 to control. Allocation of study arms was not blinded to the investigators, field team or participants. However, laboratory technicians were blinded.

### Intervention delivery

ATSB® Sarabi Bait Station (version 1.2, Westham Co., Hod-Hasharon, Israel) containing dinotefuran, a neonicotinoid insecticide, were deployed to consenting households in the 35 intervention clusters, in both core and buffer areas, from 1^st^ to 22^nd^ March 2022 (see timeline in Fig3). Two ATSB stations were hung approximately 1.8 meters above the ground on the exterior walls of all eligible household structures (those with a complete roof, walls ≥1.8 m high, and at least three complete exterior walls). The study team visited households to monitor ATSBs every two months. ATSBs that were missing or damaged were replaced, and all ATSB stations were replaced every six months. Four rounds of ATSBs were deployed over two years. All ATSBs were removed from the fieldbetween 2^nd^ April 2024 and 24^th^ April 2024.; used ATSBs were incinerated. Pyrethroid-only LLINs were distributed in Siaya county by the Kenyan Ministry of Health in June 2021, aiming to achieve universal coverage (one LLIN for every two household residents). During the trial, the study team delivered supplemental pyrethroid-only LLINs to all clusters in October 2022 to households with inadequate LLIN coverage (less than one LLIN per two residents).

### Cohort study

Three consecutive cohorts of children aged 1-<15 years were enrolled in March-April 2022, September-October 2022, and March-April 2023. The first two cohorts were followed for up to 6.5 months, while the third cohort was followed for up to 12.5 months. In total, the cohorts were followed from 7^th^ March 2022 to 7^th^ March 2024.

A list of randomly selected children aged 1-<15 years was generated for recruitment from the baseline census list for each cluster. A new list was generated for each cohort. Children on the recruitment list were invited for a screening appointment at one of nine dedicated study clinics staffed by study clinicians and community interviewers. Children were enrolled in the cohort study if they met the following selection criteria: (1) age 1 to < 15 years; (2) resident of the household (over the previous four months, and intending to stay for an additional 6-12 months); (3) provision of written informed consent by their parent or guardian; (4) provision of assent by children aged 13-14 years; (5) no evidence of pregnancy; (6) not taking cotrimoxazole prophylaxis; (7) no known sickle cell disease; (8) no contraindication to AL; and (9) not currently enrolled in another interventional study or enrolled in the previous ATSB cohort.

At enrolment, information was collected on demographics, clinical history, and self-reported LLIN use the previous night. Children were asked about history of fever within the past 48 hours, and axillary temperature was measured. A finger-prick blood sample was collected for rapid diagnostic test (RDT) for malaria (First Response® Malaria Ag pLDH/HRP2 Combo Card kit) and storage on filter paper at -80°C for future molecular studies. All participants were treated with AL, regardless of the RDT result. Two weeks after enrolment, participants were asked to return to the clinic to assess parasite clearance. Participants testing positive at the clearance visit were treated with either AL or dihydroartemisinin-piperaquine (DP), according to Ministry of Health guidelines. A household survey was conducted soon after enrolment to collect information on household characteristics, house construction, indicators of socioeconomic status, and LLIN ownership.

Cohort participants were encouraged to seek care at study clinics and were scheduled for routine clinic visits every 4 weeks. At each monthly visit, information was collected on any recent illness, care-seeking, LLIN use, and travel history. A finger-prick capillary blood sample was collected for storage on filter paper. If the cohort participant missed a scheduled visit, their parent or guardian was called. If we failed to reach them after three attempts, a field worker was sent to their house. If a participant sought care outside of the study, information on symptoms, malaria testing, and antimalarial treatment were recorded. If a child was febrile (axillary temperature *≥*37.5°C or history of fever in the previous 48 hours) at any clinic visit, a RDT was performed. If the child had been treated for malaria within the past 5 weeks, a blood smear was prepared for microscopy, instead of an RDT. If the RDT or blood smear was positive, the child was treated according to the Kenya Ministry of Health guidelines with AL or DP. Children with severe malaria were referred to the nearest hospital for treatment with parenteral artesunate. Cohort participants were excluded after enrolment if they withdrew consent, moved out of the study area, or met any of the exclusion criteria for cohort participation.

### Continuous malaria indicator surveys (cMIS)

A cMIS survey measured malaria prevalence cross-sectionally in community residents throughout the year from 5^th^ April 2022 to 11^th^ February 2024. Households from each cluster were randomly selected from a census list and were visited once. Written informed consent and assent (for children aged 13-17 years) were sought for all household residents (aged ≥1 month) who were present on the day of the survey. Information on demographics, housing characteristics, and LLIN ownership and use was obtained from the head of household, or their designee. A finger-prick blood sample was collected from each consenting participant for an RDT. Participants who tested positive were treated with AL.

### Entomology surveys

Mosquito collections using human landing catches (HLC) and CDC ultraviolet light traps (UVLT) were conducted monthly from 4^th^ April 2022 to 27^th^ March 2024 in 16 randomly selected clusters using a stratified approach to ensure study arm assignments were balanced across the area. Trained community members consented to participate as mosquito collectors. Ten households enrolled in the cohort study were randomly chosen per cluster to participate in the HLCs. Initially, HLC collections were conducted indoors and outdoors from 6:00 PM to 6:00 AM. In October 2022, the sampling period was extended to 11:00 AM to capture late morning biting. UVLT collections were paired with the HLCs and conducted in the same 16 clusters, but in ten different households located within 50 meters of each HLC household. The timing of UVLT collections mirrored the HLCs. Seven cross-sectional UVLT collections were also done in the 70 clusters, in February 2022 (baseline), May and November 2022, May, July and November 2023, and February 2024 (Fig3). At each point, ten households were randomly sampled in the remaining 54 clusters, resulting in comprehensive data from all 70 clusters.

### Laboratory procedures

For microscopy, thick and thin blood smears were prepared on the same slide by study clinicians and were transported to the laboratory in Siaya County. Details on slide preparation and microscopy procedures are included in the supplemental file. All slides were read by a second microscopist, and a third independent reviewer settled any discrepant readings. Mosquitoes collected using HLCs and UVLTs were transported to the laboratory for morphological identification. All undamaged, unfed female *Anopheles* spp. mosquitoes captured alive during HLCs were dissected to assess parity by the ovary tracheation method [22].

### Sample size calculations

Full details of the sample size calculations are provided in the trial master protocol and statistical analysis plan [14, 15]. A sample size of 1,260 person-years (630 person-years per arm) was required to detect a ≥30% reduction in the incidence of clinical malaria among children aged 1-<15 years (from 845 per 1,000 person-years in the control arm to 592 in the intervention arm over two years), assuming a coefficient of variation of 0.4, two-sided alpha of 0.049, 80% power, and an anticipated 20% loss of person-time due to loss to follow up or consent withdrawal. For cMIS, a sample size of 4,200 participants annually (60 per cluster) was required to detect a 30% reduction in RDT-confirmed malaria prevalence, from 29.0% to 20.3%, assuming an intra-cluster correlation coefficient (ICC) of 0.05, 80% power, a two-sided alpha of 0.05, and a 20% non-response rate, allowing for analysis within each 6-month sampling period. For vector parity, simulations (n=1,000) accounting for variation between clusters, household, months and monthly catch were conducted [23], assuming a two-sided alpha of 0.05, power of 80%, ICC of 0.04, expected female *Anopheles* per trap night of 2.5, and 16 clusters (8 per arm). To detect a 9%-point reduction, from 50% parity in the control arm, we estimated that ten entomological collections per cluster per month were required.

### Outcome measures

Incidence of malaria (clinical malaria cases per person-year) in children aged 1-<15 years was the primary outcome, defined as fever (axillary temperature of[]≥37.5°C or history of fever within the past 48 hours) plus a positive malaria test (RDT or microscopy). Malaria cases diagnosed within 14 days of AL treatment, or 28 days of DP treatment, were not counted as incident cases. To account for the post-treatment prophylactic effect of AL and DP, the at-risk person-time was adjusted by subtracting 14 days for each AL treatment, and 28 days for each DP treatment. Temporary absences from the study area that did not interfere with monthly clinic visits were not considered to reduce individual exposure time. However, absences >35 days were excluded from exposure time.

Secondary outcomes included (i) malaria prevalence: proportion of participants aged ≥1 month testing positive by RDT; (ii) parity: proportion of dissected mosquitoes that were parous; (iii) vector density (the number of female *Anopheles* collected per household). All entomological outcomes were calculated for all *Anopheles* spp. collected and disaggregated by species complexes based on morphological identification.

### Analytical approach

All analyses were done using the intention-to-treat population. For the primary outcome, a multi- level variance components model within a generalised linear framework with a Poisson likelihood and a log link function was used. A random intercept was included for cluster, and study arm was included as a fixed effect. We examined the distributional assumption that the mean and variance of the outcome were similar after accounting for cluster effects. Adjusted analyses including pre-specified covariates (sub-county, age, sex, housing type, baseline cluster-level malaria incidence and adequate bednet coverage, defined as one net per two residents) were conducted [15]. Other factors considered in the restricted randomisation were not included in the adjusted (or unadjusted) analyses. Household structures were classified as ‘improved’ (finished walls and roofs, and closed eaves) or ‘traditional’ (all other construction). Subgroup analyses by factors specified a priori (age, sex, baseline parasite prevalence, housing type) were also conducted by including the subgroup variable and an interaction term with study arm in an adjusted analysis. If the *p*-value of the interaction terms was <0.05, separate effect estimates and confidence intervals for each sub-group were presented.

Malaria prevalence was analysed using a multi-level variance components model within a generalised linear framework, with a Bernoulli likelihood and a logit link function. A random intercept was included for each study cluster, and the study arm was treated as a fixed effect. Adjusted analyses included pre-specified covariates (region, age, sex, bednet use) [15]. Vector parity and density were analysed using individual-level data through generalised linear mixed models. Both unadjusted and adjusted analyses were conducted, with a random effect for clusters included in the models. For the unadjusted analysis of parity, a Bernoulli likelihood was used with a logit link function and a fixed effect for the study arm. The adjusted analysis included fixed effects for collection location (indoors vs. outdoors), time since intervention, and calendar month for seasonality. For density, a negative binomial distribution was used to account for overdispersion.

The unadjusted analysis included a fixed effect for the study arm, while the adjusted model considered collection location, time since intervention, and calendar month. Analyses were completed using R version 4.4.1.

### Research ethics approval

The trial was approved by the Kenya Medical Research Institute Scientific and Ethics Review Unit (KEMRI SERU: 4189), Institutional Review Board of the US Centers for Disease Control and Prevention (IRB: 00008118), and the Liverpool School of Tropical Medicine Research Ethics Committee (LSTM REC: 21-027). A license for the trial was also granted by the National Commission for Science, Technology, and Innovation (NACOSTI) in Kenya.

## Results

### Cohort recruitment

Of 4,509 children on the cohort recruitment lists (Fig1), 804 could not be contacted (399 intervention; 405 control), and 3,705 children were screened (1,858 control; 1,847 intervention). Of these, 523 children were not eligible (250 intervention; 273 control), and 220 did not consent (111 intervention; 109 control). Reasons for exclusion are detailed in Table S1. Overall, 2,962 children were enrolled (1,497 intervention; 1,465 control), including 1,000 in cohort-1, 969 in cohort-2, and 993 in cohort-3.

### Characteristics of participants and households

Characteristics of cohort participants and households were similar in both arms (Table 1). The mean cluster-level proportion of children aged <5 years, 5-9 years, and 10-<15 years was 26.5% (SD 7.4%), 34.2% (SD 7.8%), and 39.3% (SD 8.9%), respectively. The median cluster-level prevalence of positive RDTs at enrolment was 45.7% (IQR: 28.6%, 67.9%) in the intervention arm and 48.7% (IQR: 24.3%, 61.8%) in the control. We surveyed 2,644 households of 2,886 (97%) participants. The cluster-level household size and socio-economic index were evenly distributed across both study arms. Most houses were classified as traditional construction, primarily due to open eaves. Nearly all households owned at least one bed net; however, the mean cluster-level proportion of households that owned at least one net for every two residents was only 51.4% (SD 9.7%).

**Table 1.** Cluster-level characteristics of cohort participants & households (stratified by study arm)

| Variable | Characteristic | Cohort study |  |
| --- | --- | --- | --- |
|  |  | Control arm | Intervention arm |
| Number of clusters |  | 35 | 35 |
| Number of cohort participants enrolled |  | 1,465 | 1,497 |
| Female sex, mean (SD) <sup>a</sup> |  | 47.1% (7.5%) | 48.2% (7.2%) |
| Age categories, mean (SD) <sup>a</sup> | — 1 to < 5 years | 27.5% (7.4%) | 25.4% (7.4%) |
|  | — 5 to 9 years | 32.5% (7.1%) | 35.9% (8.2%) |
|  | — 10 to < 15 years | 40.0% (8.0%) | 38.6% (9.8%) |
| RDT positive, median (IQR) <sup>a</sup> |  | 48.7% (24.3%, 61.8%) | 45.7% (28.6%, 67.9%) |
| Number of households surveyed |  | 1,311 | 1,333 |
| Household size by cluster, mean (SD) <sup>a</sup> | — [1 -4] residents | 26.4% (7.3%) | 24.7% (7.7%) |
|  | — [5 -7] residents | 54.2% (9.0%) | 56.1% (10.0%) |
|  | — 8+ residents | 17.8% (10.2%) | 17.1% (9.5%) |
| Socioeconomic index in terciles, mean (SD) <sup>a</sup> | — Poorest | 34.6% (10.7%) | 31.3% (10.5%) |
|  | — Poor | 31.5% (8.3%) | 34.3% (8.8%) |
|  | — Least poor | 33.9% (11.2%) | 34.4% (11.2%) |
| Household construction, mean (SD) <sup>a,b</sup> | — Traditional | 92.5% (5.0%) | 92.2% (4.9%) |
|  | — Modern | 7.5% (5.0%) | 7.8% (4.9%) |
| Net ownership, mean (SD) <sup>a</sup> | Households with at least one net | 98.4% (2.4%) | 98.8% (2.4%) |
| Adequate net coverage, mean (SD) <sup>a</sup> | Households with 1 net for every 2 residents | 52.0% (9.7%) | 50.8% (9.8%) |

<sup>a</sup> Proportions at the level of each cluster
<sup>b</sup> Houses were classified as ‘modern’ (synthetic walls and roofs, eaves closed or absent) or ‘traditional’ (all other construction)

### ATSB delivery

A total of 268,268 ATSBs were deployed between March 2022 and April 2024. Of the 33,419 household structures deemed eligible for ATSBs in March 2022, 183 did not consent, and another 56 did not receive ATSBs. Thus, 66,360 ATSBs were deployed to 33,180 (99.3%) structures. The mean cluster-level coverage of ATSBs was 98.7% (range 96-100%) for eligible structures. In September 2022, March 2023, and September 2023, 66,868, 67,704, 67,336 ATSBs were deployed, respectively. New structures were mapped and consented at each deployment round. Overall, during the 2- monthly monitoring visits, 12,427 (4.6%) stations were missing (not present at the time of the monitoring visit), and 42,297 (15.8%) met criteria for replacement, primarily due to mould, sugar leakage, or damage, and were replaced with new ATSBs. All stations were replaced at the 6-month mark. Further details on ATSB density and coverage will be published separately.

### Cohort follow-up

Of the 2,962 children enrolled, 2,702 (91.2%) completed the 6 follow-up visit (1,375 intervention; 1,327 control). In cohort-3, 876 (88.2%) of 993 children completed the 12 follow-up visit (448 intervention; 428 control). Overall, 2,967 routine and sick visits (1,525 intervention; 1,442 control) were associated with fever (elevated temperature ≥ 37.5⁰C or history of fever in the past 48h). In the intervention arm, 937 (71.5%) of 1,310 children tested with an RDT, and 131 (59.6%) of 220 children tested by microscopy, were positive. In the control arm, 821 (66.4%) of 1,236 children tested with an RDT, and 110 (49.8%) of 221 children tested by microscopy, were positive. Overall, 2,869 (96.9%) participants (1,461 intervention, 1,408 control) were included in the primary endpoint analysis (Fig1). Children were excluded if they missed the clearance visit, lacked a microscopy result at the clearance visit (26 intervention; 37 control), or did not accrue eligible person-time after clearance (9 intervention; 18 control).

### Impact on primary outcome: clinical malaria incidence

We captured 1,829 incident malaria cases over 1,445 person-years of follow-up. Malaria incidence was 1.32 episodes per person-years in the intervention versus 1.20 in the control arm (Table 2).

**Table 2.**
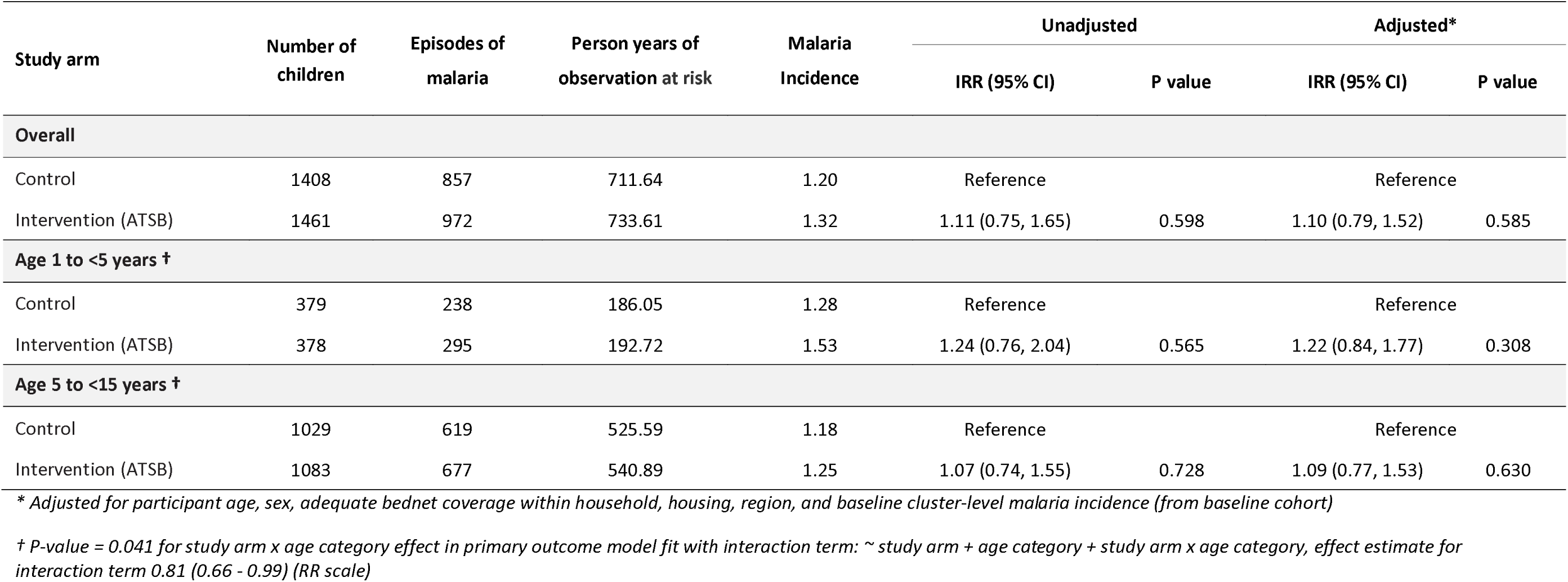
Malaria incidence (primary outcome)

| Study arm | Number of children | Episodes of malaria | Person years of observation at risk | Malaria Incidence | Unadjusted |  | Adjusted* |  |
| --- | --- | --- | --- | --- | --- | --- | --- | --- |
|  |  |  |  |  | IRR (95% CI) | P value | IRR (95% CI) | P value |
| Overall |  |  |  |  |  |  |  |  |
| Control | 1408 | 857 | 711.64 | 1.20 | Reference |  | Reference |  |
| Intervention (ATSB) | 1461 | 972 | 733.61 | 1.32 | 1.11 (0.75, 1.65) | 0.598 | 1.10 (0.79, 1.52) | 0.585 |
| Age 1 to <5 years † |  |  |  |  |  |  |  |  |
| Control | 379 | 238 | 186.05 | 1.28 | Reference |  | Reference |  |
| Intervention (ATSB) | 378 | 295 | 192.72 | 1.53 | 1.24 (0.76, 2.04) | 0.565 | 1.22 (0.84, 1.77) | 0.308 |
| Age 5 to <15 years † |  |  |  |  |  |  |  |  |
| Control | 1029 | 619 | 525.59 | 1.18 | Reference |  | Reference |  |
| Intervention (ATSB) | 1083 | 677 | 540.89 | 1.25 | 1.07 (0.74, 1.55) | 0.728 | 1.09 (0.77, 1.53) | 0.630 |

\* Adjusted for participant age, sex, adequate bednet coverage within household, housing, region, and baseline cluster-level malaria incidence (from baseline cohort)
† P-value = 0.041 for study arm x age category effect in primary outcome model fit with interaction term: ~ study arm + age category + study arm x age category, effect estimate for interaction term 0.81 (0.66 - 0.99) (RR scale)

There was no evidence of a difference in malaria incidence between the intervention and control group in the unadjusted analysis (incidence rate ratio [IRR] 1.11; 95% CI: 0.75-1.65; p=0.598) or in the multivariable analysis. The median cluster-level incidence of malaria overall was 1.275 cases per person-year (range 0.04 to 4.59 cases per person-year). The coefficient of variation for malaria incidence in the trial was 0.75. Subgroup analysis suggested that the effect of ATSBs varied by age (1-<5 years: adjusted IRR 1.22 [95% CI: 0.84-1.77]; 5-<15 years: adjusted IRR 1.09 [95% CI: 0.77-1.53], p-value for study arm x age group interaction=0.041) (Table 2). There were no sub-group differences for other pre-specified factors. Overall, 18 serious adverse events (9 intervention; 9 control) were reported; all were considered unrelated to the ATSB intervention (Table S2).

### Impact on secondary outcomes: malaria prevalence

In the cMIS survey, 7,488 community residents (3,760 intervention; 3,728 control) from 1,685 households (850 intervention; 835 control) were tested for malaria using RDTs (Table 3). Of these, 1,474 (39.2%) intervention and 1,461 (39.2%) control participants tested positive. There was no difference in the odds of malaria infection between the two groups in the unadjusted analysis (odds ratio [OR] 0.98; 95% CI: 0.60-1.59; p-value 0.93), or in the multivariable analysis (Table 3).

**Table 3.** Secondary epidemiological and entomological outcomes.

| Survey | Indicator |  |  | Study arm | Unadjusted |  |  | Adjusted |  |  |
| --- | --- | --- | --- | --- | --- | --- | --- | --- | --- | --- |
|  |  |  |  |  | n/N (%) | OR [95% CI] | p-value | OR [95% CI] | p-value |  |
| Continuous malaria indicator survey | RDT positivity (all ages) * |  |  | Control | 1461/3728 (39.19) | Ref |  | Ref |  |  |
|  |  |  |  | Intervention (ATSB) | 1474/3760 (39.20) | 0.98 (0.60–1.59) | 0.93 | 0.97 (0.68–1.37) | 0.85 |  |
| Entomology surveillance | Parity (Female Anopheles spp.) † |  |  | Control | 1973/2399 (82.2) | Ref |  | Ref |  |  |
|  |  |  |  | Intervention (ATSB) | 3579/4058 (88.2) | 1.34 (0.905–1.99) | 0.14 | 1.30 (0.87–1.94) | 0.201 |  |
| Indicator | Study arm | Households sampled | Total Collected | Household Mean (SD) | Household Median (IQR) | Cluster Median (IQR) | IRR (95% CI) | p-value | IRR (95% CI) | p-value |
| Density by HLCs | Control | 1925 | 10,846 | 5.63 (13.6) | 0 (0 – 4) | 2.06 (1.20–7.52) | Ref |  | Ref |  |
| (Female *Anopheles* spp.) | Intervention | 1924 | 28,840 | 15 (46) | 0 (0 – 5) | 0.913 (0.52–11.2) | 0.954 (0.145–6.27) | 0.96 | 0.985 (0.142–6.82) | 0.99 |
| Density by UVLTs | Control | 1927 | 18,191 | 9.44 (22.1) | 3 (0 – 9) | 5.46 (3.93–10.8) | Ref |  | Ref |  |
| (Female *Anopheles* spp.) | Intervention | 1924 | 37,531 | 19.5 (52.7) | 2 (0 – 14) | 4.08 (1.96–22.5) | 0.960 (0.257–3.59) | 0.952 | 0.94 (0.244–3.63) | 0.929 |
| Density by UVLT CSS | Control | 2103 | 22,797 | 10.8 (21.7) | 3 (0 – 12) | 7.15 (3.78–13.6) | Ref |  | Ref |  |
| (Female *Anopheles* spp.) | Intervention | 2104 | 29,792 | 14.2 (28.4) | 3 (0 – 15) | 5.58 (2.71–20.3) | 1.02 (0.548–1.90) | 0.952 | 0.954 (0.63–1.44) | 0.822 |

\* cMIS analysis adjusted for study arm (ATSB), region, sex, age in year of survey & bednet use (slept under net last night)
† Parity analysis includes fixed effects for collection location (indoors vs. outdoors), time since intervention and calendar month as a seasonality adjustment
‡ Adjusted analysis for density includes fixed effects for study arm (ATSB) + log(baseline anopheline species number) and random effects for cluster & round

### Impact on secondary outcomes: entomology indicators

Between April 2022 and March 2024, 6,457 female Anopheles spp were collected and evaluated for parity (4,058 intervention; 2,399 control) (Table 3). Of these, 3,579 (88.2%) in the intervention arm and 1,973 (82.2%) in the control arm were parous. There was no difference in the odds of parity between the groups in the unadjusted analysis (OR 1.34; 95% CI: 0.905-1.99; p=0.14), or in the multivariable analysis. Similarly, no evidence of difference in the density of female Anopheles spp was found when using human landing catches or UV light traps in the unadjusted or adjusted analyses. Species-specific results for An. funestus and An. gambiae were similar, with no significant differences in parity or density between the intervention and control groups stratified by species or collection strategy (Table S3). Detailed results of the entomology surveys will be published separately.

## Discussion

In this trial, we evaluated the impact of ATSBs on malaria indicators in western Kenya. Despite promising results from a preliminary entomology study in Mali [10], epidemiological modelling suggesting that ATSBs could substantially reduce malaria burden in similar settings [11], and a choice experiment in semi-field enclosures in Kenya suggesting that ATSBs were more attractive to local Anopheles mosquitoes than natural sugar sources [12], we found no evidence that ATSBs reduced malaria incidence in children aged 1-<15 years, malaria prevalence in community residents of all ages, or vector parity or density in western Kenya. Our results are consistent with the findings from the partner trial in Zambia, which found no impact of ATSBs on clinical malaria incidence, parasite prevalence or entomological indicators in a setting with seasonal malaria transmission [24, 25]. The lack of ATSB effectiveness is likely attributable to multiple factors, including competing sugar sources for vector feeding in Kenya [26, 27], uncertainty about optimal intervention coverage in this setting, product design and limited interaction with the product by local vector populations [10, 28]. For ATSBs to effectively control malaria, a sufficient proportion of the mosquito population must be attracted to, subsequently feed on the insecticide-laced syrup in ATSB stations, and be killed, thus reducing the vector population and ultimately malaria transmission. It is unclear where along this pathway ATSBs failed in Kenya. However, the lack of entomological impact on vector density suggests that ATSBs did not work in western Kenya, as they appeared to in the Mali trial [10].

Why were ATSBs not effective in Kenya? Did ATSBs fail to lure mosquitoes, perhaps due to competition with other natural sugar sources? This seems plausible given the range and quantity of natural sugar sources in the environment which could lead to insufficient coverage [29]. Did mosquitoes fail to feed on the ATSBs, or did the insecticide fail to kill mosquitoes? This is possible, although laboratory experiments suggest that the mosquitoes did feed on the devices, and those that fed, died [30]. Or, was the proportion of mosquitoes affected by ATSBs insufficient to reduce vector density, parity and transmission in our setting? This seems most likely. Inadequate density and coverage of ATSB devices, the abundance of alternative food sources, or the sheer numbers of mosquitoes in the area may have all contributed. Modelling studies on mosquito feeding behaviour and the potential of ATSBs to control mosquito populations and Plasmodium falciparum malaria prevalence were fitted to data from Mali [11, 13]. However, natural sugar sources are more abundant in western Kenya than in Mali. The effectiveness of ATSBs may be context specific with higher efficacy in more arid regions. Studies in Mali and Israel, sites with less natural sugar sources, have demonstrated a strong entomological impact of ATSBs [10, 26, 31, 32]. However, in Zambia, the Phase III cluster-randomised trial evaluating ATSBs found that while vector density was reduced (but not significantly) by 35%, no difference was observed in parity, sporozoite rate or EIR [24], and correspondingly no significant reductions in clinical malaria incidence or malaria prevalence were observed [25]. Full results of the companion trial in Mali may shed more light on the efficacy of ATSBs in different settings.

Our study had several limitations. First, we currently lack a specific density target for spatial deployment of ATSBs, and it remains unclear how bait stations interact with environmental factors such as vector larval site locations, natural sugar sources, housing construction and human settlement patterns. Further research is needed to better understand the impact of intervention density and coverage, to optimise their effectiveness in diverse malaria transmission settings.

Second, the calculated coefficient of variation for malaria incidence, which measures heterogeneity between clusters, was 0.75 in the trial, higher than the estimate of 0.4 included in our sample size calculations. Underestimating coefficient of variation can compromise trial power and reduce effect size precision in cluster-randomised trials [33]. However, all outcomes measured in this trial (malaria incidence and prevalence, vector parity and density) were greater in the intervention arm than in the control, suggesting that we were unlikely to have rejected the null hypothesis in the trial, even if it had been more sufficiently powered to detect a statistically significant reduction in malaria incidence.

## Conclusion

In a high transmission area in western Kenya, we found no evidence that ATSBs reduced malaria incidence, prevalence, or vector parity and density, consistent with the results from the companion trial in Zambia. Results from the partner trial Mali are expected soon, which may provide more insight into the effectiveness of ATSBs in different transmission settings. Future work on ATSBs could focus on steps in the causal pathway of impact (attraction, feeding, and mortality), the complex interplay between sugar feeding and host seeking by malaria vectors, and density and coverage of ATSBs in a variety of environmental and transmission settings. Further research on innovative malaria control interventions is also needed to address outdoor biting and residual transmission, and to help build the toolbox for integrated vector management.

## Data Availability

De-identified data are available from the corresponding author on reasonable request. Following publication of forthcoming secondary analyses of trial data, the de-identified trial dataset will be posted on a public repository.

## Acknowledgments

We would like to thank the clinicians, monitors, community interviewers, human landing catch volunteers, the entomology laboratory and field teams, and community health promoters for their dedicated efforts in the collection of data and ensuring the smooth running of study activities. We are grateful to the social science team led by George Okello and Teresa Bange for helping us develop our community engagement strategy. We extend our appreciation to the Siaya County Ministry of Health for approving this study. We also thank the sub-county health management teams (Alego- Usonga and Rarieda) for facilitating community engagement and the health facility in-charges for their assistance with the cohort study and allowing their facilities to be used for storage of study supplies. Special thanks to Timothy Orwa the community liaison who was instrumental in community sensitization, Benta Kamire, Helen Wong, Sheila Nyarinda, and Maurice Ondeng for their instrumental roles in facilitating study activities, Dickens Riaga for ensuring the internal consistency of the trial and Dennis Omondi, the KEMRI/ CGHR Siaya laboratory manager for overseeing sample management. We are grateful to the Siaya County Commissioner and the national government staff who enabled us to conduct field activities without any issues. Lastly, we are grateful to all participants for their willingness to take part in this trial.

## Author’s contributions

EO, FtK, and AMS conceived the trial, with input from DM, JJ, MJD, and SK. EO, FtK, and AMS designed the study, developed the procedures, and drafted the protocol with contributions from DM, ML, JJ, CO, MC, MM, OT, WO, KO1, BS, SO, JK, and BP. CO, KO1, BS, SO, JK, and BP led field activities and data collection, under the oversight of SGS, FtK, AMS, and EO, with support from JS, JRG, JEG, and MJD. AK, DM, FA, and OT managed the data, and ML led the data analysis, with support from AK, DM and SGS. SO and JK led the entomology surveillance and mosquito sample processing. KO2 coordinated the collection, processing, and storage of blood samples. CO and SGS interpreted the data and drafted the manuscript, with input from ML, DM, and AK. All authors reviewed the manuscript and approved it for publication. CO, the corresponding author, had full access to all the data in the study and held final responsibility for the decision to submit the manuscript for publication.

## Supplemental files

S1. Malaria microscopy procedures

Table S1. Cohort study recruitment

Table S2. Serious adverse events & deaths

Table S3. Additional parity outcomes

---

Fig 1: CONSORT flow diagram

Fig 2: Map of study area: The study included 70 clusters, each consisting of a minimum of 100 households in the core area. Clusters were randomised 1:1 to intervention (ATSB) or control (standard care). A sub-set of 16 clusters were randomly selected for entomology surveillance.

Fig 3: Study schema. Timeline of study activities between February 2022 to March 2024 Abbreviations – attractive targeted sugar bait (ATSB), continuous malaria indicator survey (cMIS), human landing catches (HLCs), ultraviolet light traps (UVLTs), cross-sectional surveys (CSS). Activities featured, include: four rounds of ATSB deployment and monitoring; enrolment and follow-up of three consecutive cohorts of children; two years of cMIS monitoring; seven cross-sectional mosquito collections using UVLTs; one year of entomology surveillance using HLCs and UVLTs for mosquito collections.

